# Multi-trait Analysis of GWAS for circulating FGF23 Identifies Novel Network Interactions Between HRG-HMGB1 and Cardiac Disease in CKD

**DOI:** 10.1101/2024.03.04.24303051

**Authors:** Farzana Perwad, Elvis A. Akwo, Nicholas Vartanian, Larrry J Suva, Peter A Friedman, Cassianne Robinson-Cohen

## Abstract

**Background:** Genome-wide association studies (GWAS) have identified numerous genetic loci associated with mineral metabolism (MM) markers but have exclusively focused on single-trait analysis. In this study, we performed a multi-trait analysis of GWAS (MTAG) of MM, exploring overlapping genetic architecture between traits, to identify novel genetic associations for fibroblast growth factor 23 (FGF23).

**Methods:** We applied MTAG to genetic variants common to GWAS of 5 genetically correlated MM markers (calcium, phosphorus, FGF23, 25-hydroxyvitamin D (25(OH)D) and parathyroid hormone (PTH)) in European-ancestry subjects. We integrated information from UKBioBank GWAS for blood levels for phosphate, 25(OH)D and calcium (n=366,484), and CHARGE GWAS for PTH (n=29,155) and FGF23 (n=16,624). We then used functional genomics to model interactive and dynamic networks to identify novel associations between genetic traits and circulating FGF23.

**Results:** MTAG increased the effective sample size for all MM markers to n=50,325 for FGF23. After clumping, MTAG identified independent genome-wide significant SNPs for all traits, including 62 loci for FGF23. Many of these loci have not been previously reported in single-trait analyses. Through functional genomics we identified Histidine-rich glycoprotein (*HRG*) and high mobility group box 1(*HMGB1*) genes as master regulators of downstream canonical pathways associated with FGF23. HRG-HMGB1 network interactions were also highly enriched in left ventricular heart tissue of a cohort of deceased hemodialysis patients.

**Conclusion:** Our findings highlight the importance of MTAG analysis of MM markers to boost the number of genome-wide significant loci for FGF23 to identify novel genetic traits. Functional genomics revealed novel networks that inform unique cellular functions and identified *HRG-HMGB1* as key master regulators of FGF23 and cardiovascular disease in CKD. Future studies will provide a deeper understanding of genetic signatures associated with FGF23 and its role in health and disease.

## INTRODUCTION

Mineral bone disorder related to chronic kidney disease (CKD-MBD) leads to impaired skeletal and cardiovascular homeostasis and is associated with higher fracture risk, vascular calcification, and cardiovascular-related morbidity and mortality [1]. Therapeutic strategies to prevent and treat CKD-MBD include maintenance of optimal circulating concentrations of mineral markers (MM) including parathyroid hormone (PTH), calcium, phosphate, and 25-hydroxyvitamin D and fibroblast growth factor 23 (FGF23). However, therapeutic advances have not led to meaningful decrease in morbidity and mortality rates related to CKD-MBD.

We and others have shown that genome-wide association studies (GWAS) in the general population and in CKD have identified common variants associated with circulating calcium, phosphate, PTH, vitamin D and FGF23 [2–6]. In addition, Mendelian randomization studies that leverages genetic determinants of a risk factor to determine causality have shown that genetic predictors of FGF23 excess was associated with higher heart failure risk in those with genetically predicted lower GFR [7]. However, previous studies have exclusively focused on single-trait analysis leaving gaps in comprehensive understanding of genetic drivers of CKD-MBD. In this study, we performed a multi-trait analysis of GWAS (MTAG) data of MM markers, exploring overlapping genetic architecture between the traits, to identify novel genetic associations for FGF23. We then performed a series of integrated network analyses to re-prioritize quantitative genetic traits and build relevant biological network models that identify core disease-associated genes. Through this omnigenic approach we have identified previously unknown genetic traits associated with novel biological networks at the cellular and tissue level that play a major role in cardiiovascular disease in CKD.

## METHODS

### Multi-trait analysis of GWAS

We conducted multi-trait analysis of genome-wide association studies (MTAG) of five genetically correlated mineral metabolism markers (serum FGF23, PTH, phosphate, calcium, and vitamin D) using summary-level data from 5 large-scale discovery GWAS. GWAS data for FGF23 and PTH were from 16,624 and 29,155 individuals of European ancestry in the Cohorts for Heart and Aging Research in Genetic Epidemiology (CHARGE) consortium [8], and summary-level GWAS of serum phosphate, 25(OH)D and calcium (n=366,484) were obtained from the UK Biobank [9].

Details of the MTAG approach have been described elsewhere [10]. Briefly, this approach leverages the genetic correlation between related traits to perform joint genome-wide analyses of multiple traits hence augmenting the available statistical power to detect novel genetic signals for each trait analyzed. As an extension of the inverse-variance meta-analysis in the multi-trait setting, the MTAG estimator uses single-trait GWAS summary statistics as inputs and generates trait-specific SNP effects and P-values. Beyond the advantage of requiring only GWAS summary statistics, the MTAG estimator adequately accounts for potential sample overlap in the GWAS of the different traits included in the analysis using bivariate linkage disequilibrium (LD) score regression of each pair of traits.

#### Datasets

Genomic data deposited in NCBI (GSE160145) from explanted whole human hearts from the CAIN (Cardiac Aging IN CKD) Cohort were used for our comparison analysis. Details of CAIN study were previously described [11]. In brief, hearts from deceased donors >18 yrs of age with end stage kidney disease on hemodialysis (n=13) and healthy control subjects (n=5) who had no significant comorbidities were collected post mortem to conduct a comprehensive histopathologic and genomic analysis of the left ventricle. For our comparison analysis of network interactions associated with MTAG_FGF23, we used published genomic data from bulk RNA sequencing of left ventricular cardiac tissue obtained from the CAIN cohort.

### Functional Enrichment Analysis and Gene Ontology

To delineate the potential biological implications of the genes in the MTAG_FGF23 dataset, we explored Gene Ontology (GO) terms and Kyoto Encyclopedia of Genes and Genomes Pathway Enrichment (KEGG) pathways [12–15]. The three categories of biological process (BP), molecular function (MF), and cellular component (CC) constituted the GO term. GO and KEGG terms with a false discovery rate (FDR) of less than 0.05, were considered significant. The top 150 genetic traits from MTAG_FGF23 were used as input data. Using either BP, CC, or MF analysis as a baseline, the top 5 terms were selected and further annotated and visualized. KEGG enrichment analysis results are represented visually using a flowchart.

### Pathway analysis

We performed gene-based variant effect analysis considering direct and indirect relationships with IPA software (IPA®, QIAGEN Redwood City) to evaluate prior knowledge existing within IPA knowledge base and from published literature. This method maps over-representation of the top 150 MTAG_FGF23 genetic loci in various disease and functional pathways through SVM learning. To determine the top biological functions associated with MTAG_FGF23, we performed a downstream effect analysis from which we extracted the most significantly affected functions (p-value ≤ 0.05; z-score ≥ 2). The right-tailed Fisher’s exact test was used to estimate the probability that association between a set of molecules and a function or pathway might be due to random chance. The IPA z-score algorithm was used to predict the direction of change for a given function. A z-score ≥ 2 implies function is significantly increased/activated whereas a z-score ≤ 2 indicates a significantly decreased/inhibited function. We selected the most statistically significant and enriched pathways and networks to display using IPA’s visualization tools. We then performed a comparison analysis of the enriched pathways within our dataset with published cardiac RNAseq data from the CAIN cohort described above.

### Network-wide association study (NetWAS)

NetWAS is a support vector machine (SVM) learning approach that trains a classifier through grouped disease-associated genes [16]. The SVM classifier locates an optimal hyperplane from the high-dimensional predictor space to separate positive genes from negative genes. The classifier is constructed using a tissue network relevant to a disease, where the features of the classifier are the edge weights of the labeled examples to all the genes in the network. Genes are re-ranked using their distance from the hyperplane, A score is then assigned to each gene, representing the distance from the gene to the hyperplane. Genes are re-ranked using their distance from the hyperplane, which represent a network-based prioritization of a GWAS. Genes that score higher are more likely to be disease related. We used MTAG_FGF23 variants as input data after conversion of the associated P value of SNP level to gene level via the versatile gene-based association study (VEGAS) web platform [17]. Genes were reprioritized and results were ranked according to their scores to create tissue-specific gene clusters for kidney, bone and heart. Genes with high scores and that obtained consistent results in sensitivity analysis were considered to be potential causal genes.

### Statistical analyses

We performed the MTAG analysis of the single-trait GWAS of 5 mineral metabolism markers with a focus on FGF23 as the primary analysis, using the conventional Python command line tool. The MTAG estimator is an efficient generalized method of moments estimator that generates trait-specific SNP effects by performing a weighted sum of the GWAS estimates while accounting for the correlation in true SNP effects and the correlation in their estimation error (related to phenotypic correlation with sample overlap and correlated biases in SNP effects). The stepwise algorithm estimates a variance-covariance matrix of the SNPs’ estimation error (using univariate and bivariate LD score regression), the variance-covariance matrix of the SNP effects (assumed to be homogeneous across all SNPs) and then computes individual SNP effects across all traits using a closed-form solution. Independent genetic signals for FGF23 were obtained by clumping genome-wide significant signals at an r^2^ of 0.01 in 500kb windows. We probed the credibility of our findings in our work, in part by computing the false discovery rate for each SNP.

## RESULTS

### General Characterization

#### FGF23 MTAG

The strength of genetic correlations is important for investigating the pleiotropic effect of genetic variants on traits. Multi-trait analyses augments the statistical power to detect novel genetic signals to yield plausible and replicable results. We performed MTAG analysis after combining GWAS of FGF23, PTH, 25(OH)D, phosphorus, and calcium and identified 62 genomic regions for FGF23, 57 loci were novel (not within ±500kb of previously known loci) (Figure 1, Table 1). The top locus was near TKT (rs73188394), where every additional copy of the G allele was associated with a 21% higher circulating FGF23 (p=8.6 x 10^-118^).

**Figure 1.**
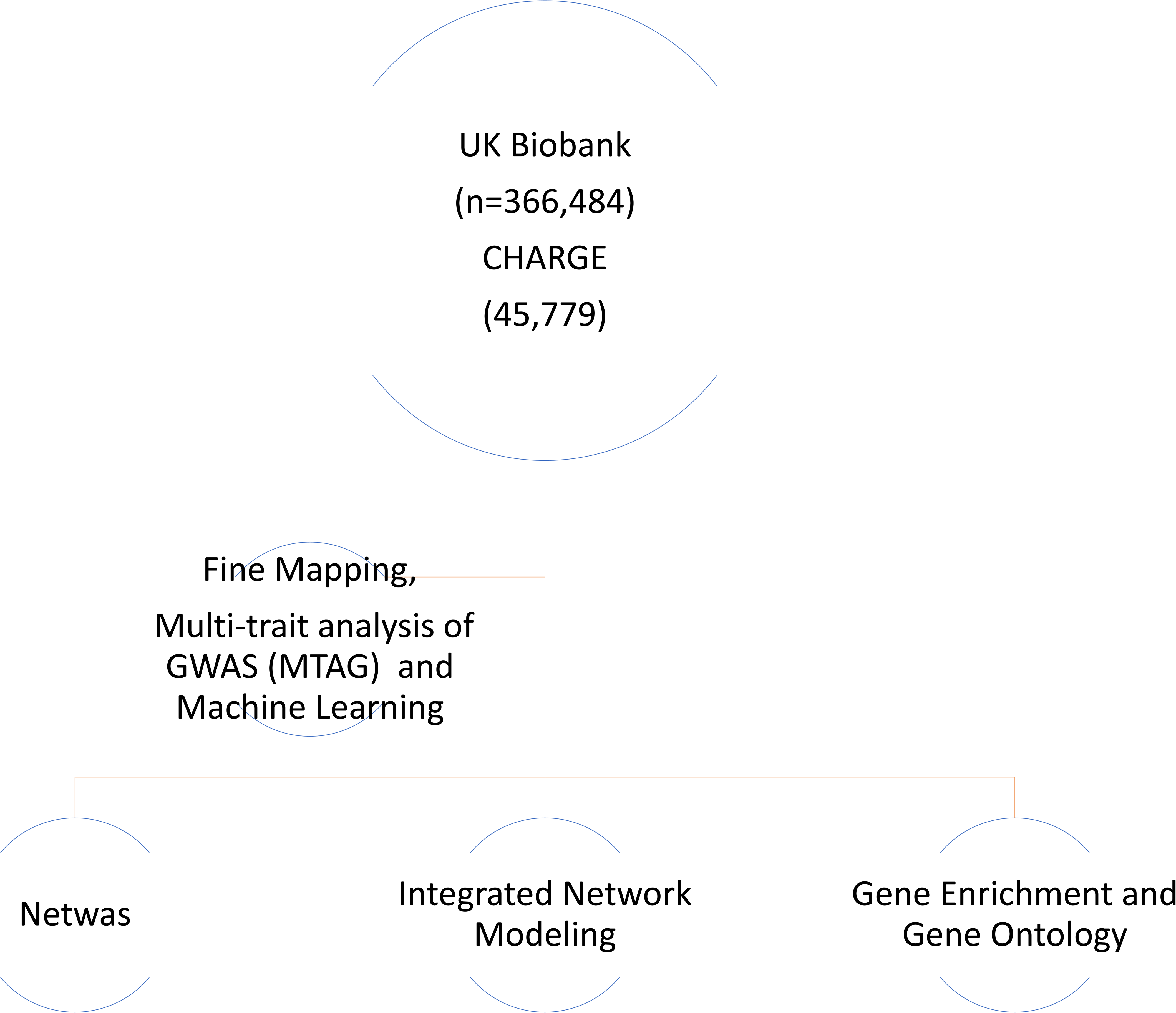
Overall Study Design

**Figure 1.**
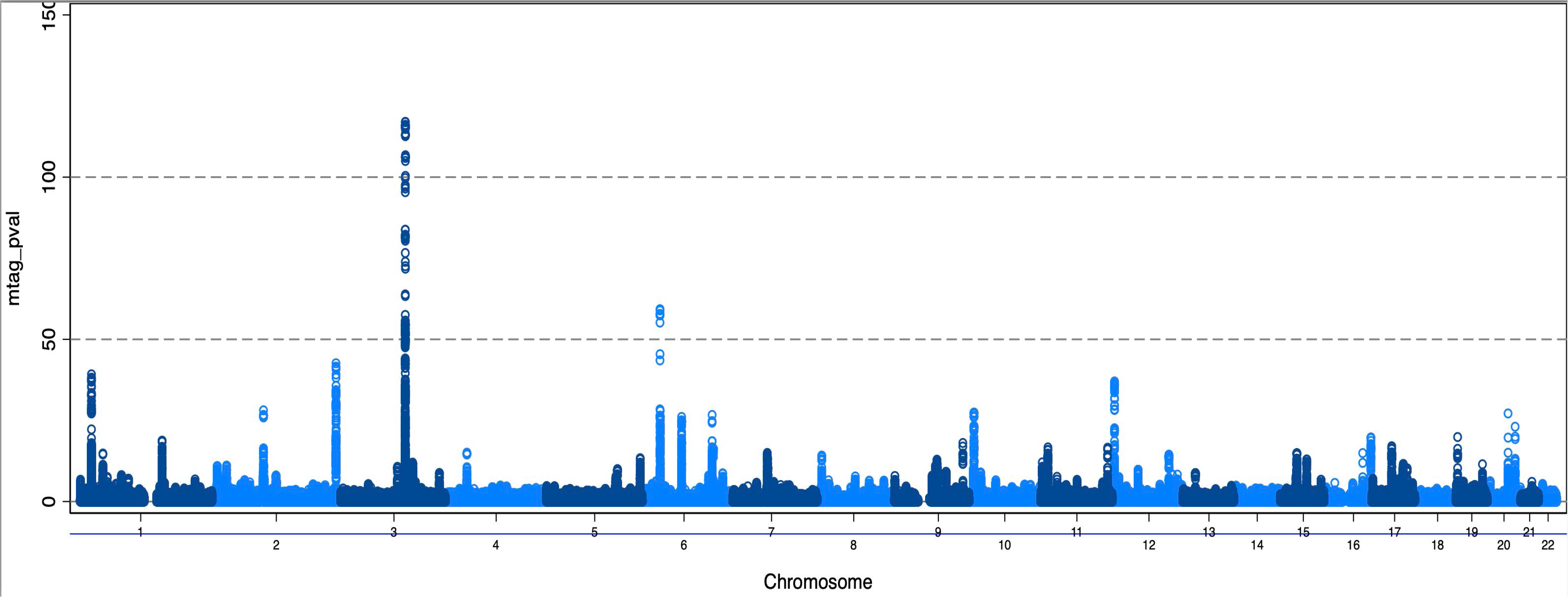
Plot shows 62 loci from FGF23 MTAG. In these plots, the y axis shows the P values of SNPs in log–log scale. The red horizontal line is the genome-wide significance level at P = 5 × 10^−8^. SNPs with P < 1 × 10^−4^ are not shown in the plots.

**Table 1.**
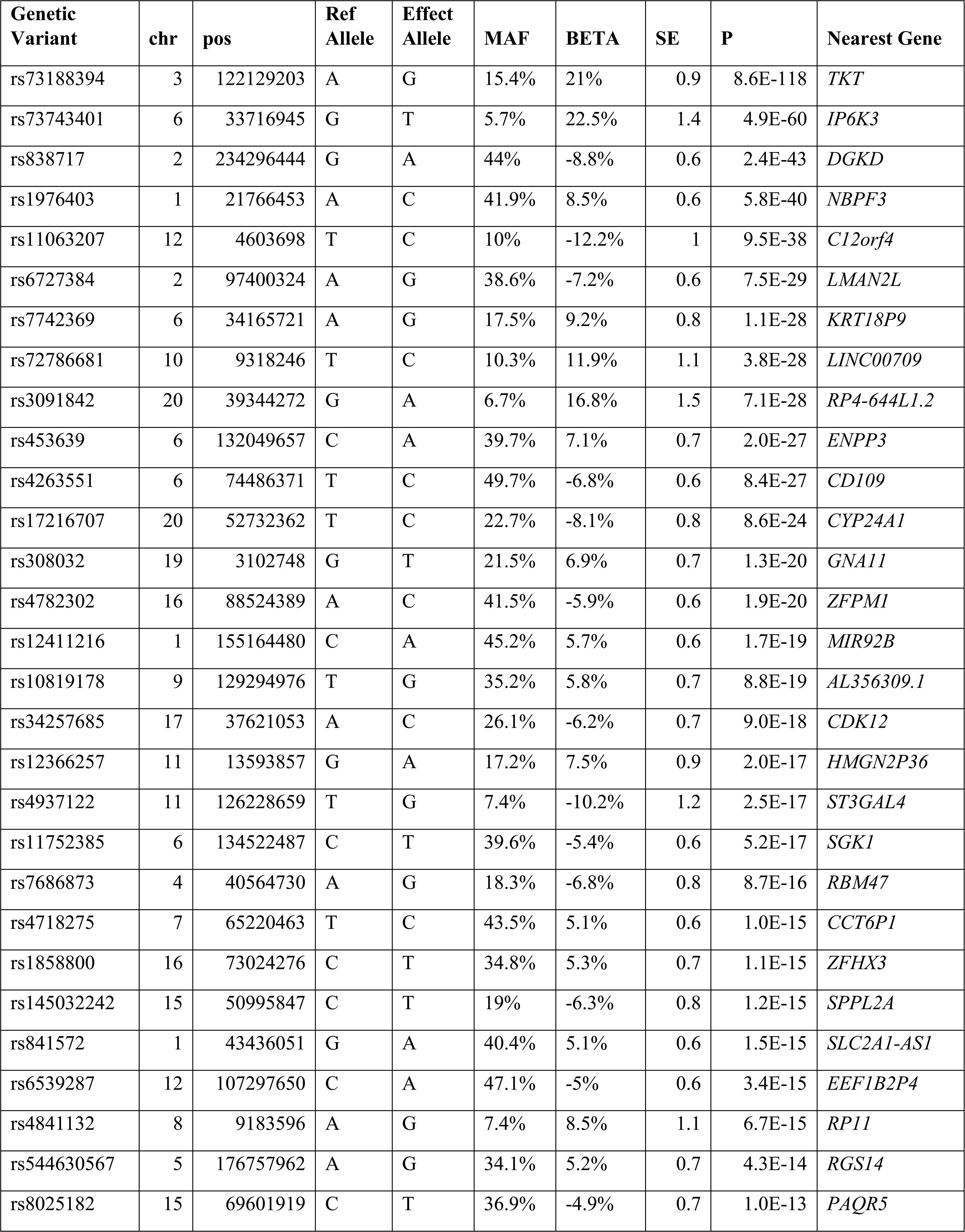
Top 30 independent genetic loci associated with circulating FGF23 from MTAG analyses.

**Table 2.** Canonical Pathway for CLEAR signaling.

| Symbol | Entrez Gene Name | SNP variant | Beta coefficient | p-value | Location | Type(s) |
| --- | --- | --- | --- | --- | --- | --- |
| *FNIP1* | folliculin interacting protein 1 | rs11950815 | 3.66 | 1.63E-08 | Cytoplasm | other |
| *GBA1* | glucosylceramidase beta 1 | rs12411216 | 5.71 | 8.69E-12 | Cytoplasm | enzyme |
| *GUSB* | glucuronidase beta | rs4718275 | 5.08 | 9.49E-38 | Cytoplasm | enzyme |
| *ITPR3* | inositol 1,4,5-trisphosphate receptor type 3 | rs73743401 | 22.45 | 2.25E-08 | Cytoplasm | ion channel |
| *PPP2R3A* | protein phosphatase 2 regulatory subunit B"alpha | rs1393787 | 5.05 | 9.49E-38 | Nucleus | phosphatase |
| *RALB* | RAS like proto-oncogene B | rs6542581 | -3.91 | 1.28E-20 | Cytoplasm | enzyme |

#### Network-wide association study (NetWAS)

Although GWAS has transformed our understanding of complex human diseases like CKD, only a small fraction of total heritability can be explained by disease-associated variants identified by GWAS. To address this “missing heritability” we used NetWAS to identify the cumulative weak effects of variants, many of which fall below statistical significance. Netwas integrates tissue-specific networks with GWAS and has an advantage of mitigating literature bias. Thus, candidate genes which are not well represented in the current literature but exhibit strong support for pathogenesis can be discovered agnostically [18]. We thus tested the omnigenic hypothesis which describes that genes can affect each other through their tightly interconnected networks, and as such, genes that have little direct bearing on a particular disease may, in aggregate, affect core disease pathways and influence disease risk. Using this approach we identified significant gene clusters in kidney, bone and heart using MTAG data. We found 9 distinct gene clusters in kidney and bone and 6 gene clusters in the heart including modules that regulate mineral and ion homeostasis. **(Figure 3A)** (A full list of gene clusters are available upon request). Given the intricate relationship between the 3 organ systems in the pathophysiology of CKD-MBD, we examined the modules for overlapping genes and found 533 genes were common in the netwas module discovery network **(Figure 3B)**. The overlapping genes were subjected to further network analyses and we found aryl hydrocarbon signaling with estrogen receptor 2 as the top canonical pathway common to all 3 organ systems. In addition, HNF4α was identified as a top upstream regulator and signaling by HIFα, TGFβ, mitochondrial dysfunction, Nrf2-mediated oxidative stress response and PPAR α/RXR α were among the top canonical pathways **(Figure 3C)**.

**Figure 3.**
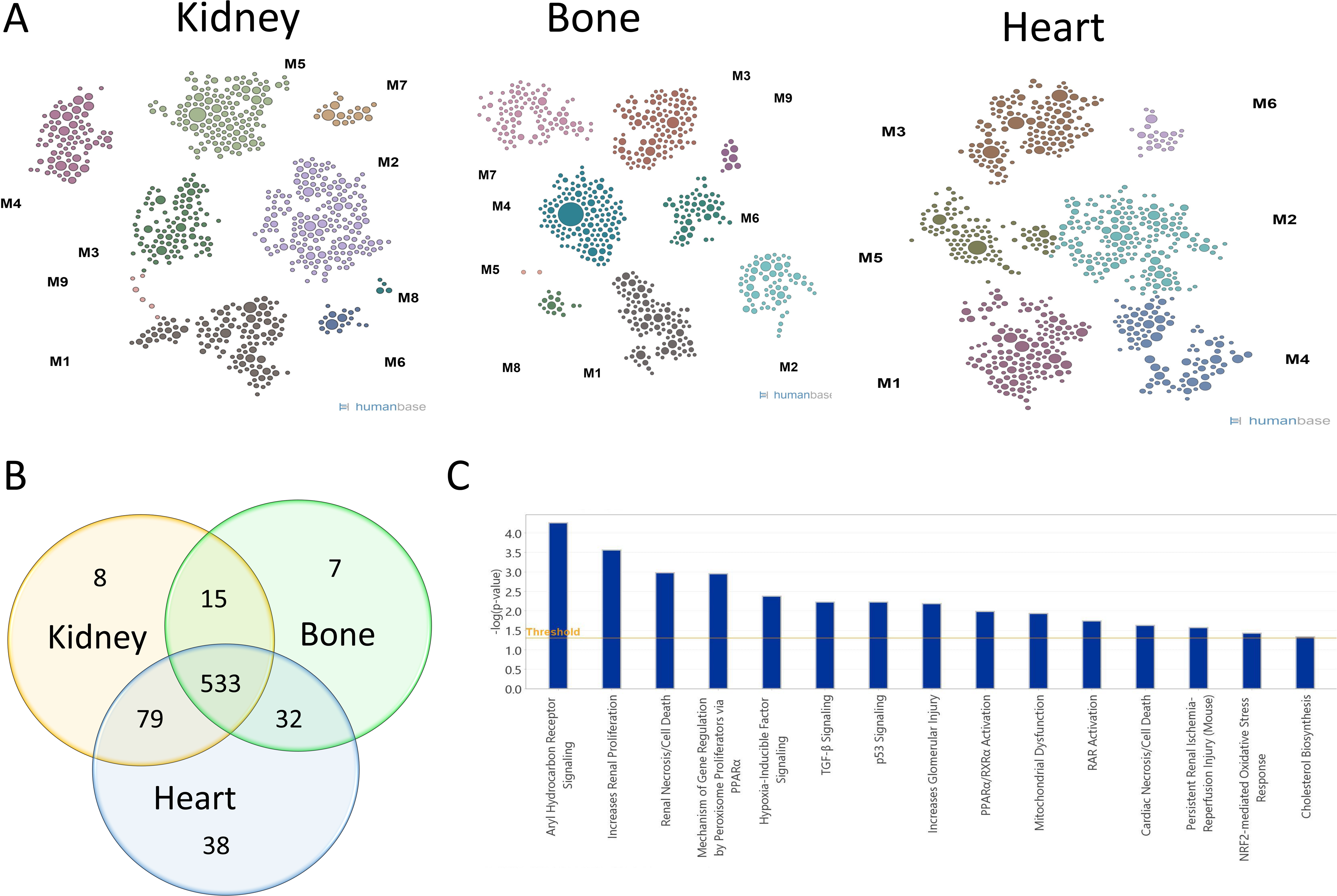
NetWAS: A Tissue-specific network-based functional interpretation of gene variants from MTAG. **A** Module discovery network at tissue level for kidney, bone and heart. **B.** Venn diagram for overlapping genes in the NetWAS modules. **C.** Top canonical pathways common to kidney, bone and heart tissues for the NetWAS gene clusters.

#### Functional Enrichment Analysis

We further explored biological implications of the variants in MTAG_FGF23 with GO and KEGG enrichment analyses. GO term “*biological processes”* was enriched for 2 key pathways; skeletal development and vitamin D **(Figure 4)** which confirms the biological relevance of MTAG variants given FGF23 is a potent regulator of vitamin D metabolism and skeletal mineralization [19–21]. We also queried the top genes from Table 1 with KEGG pathway analysis and found metabolic pathways for pentose phosphate, and phosphatidyl inositol signaling were highly enriched **(Figure 5 and 6)**. Genes within the pentose phosphate pathway were also found in the overlapping gene clusters for kidney, bone and heart in the NetWAS analyses described above. Interestingly, we found the top SNP, rs73188394, was in proximity to Transketolase *(TKT)* gene that belongs to the pentose phosphate pathway and was the most strongly associated SNP in the MTAG. Transketolase enzyme catalyzes reactions in the non oxidative component of pentose phosphate pathway and plays a critical role in energy metabolism and oxidative stress response. Pentose phosphate pathway is highly conserved in evolutionary biology, and metal ions including iron and calcium, are vital for the non enzymatic reactions in this pathway [22].

**Figure 4.**
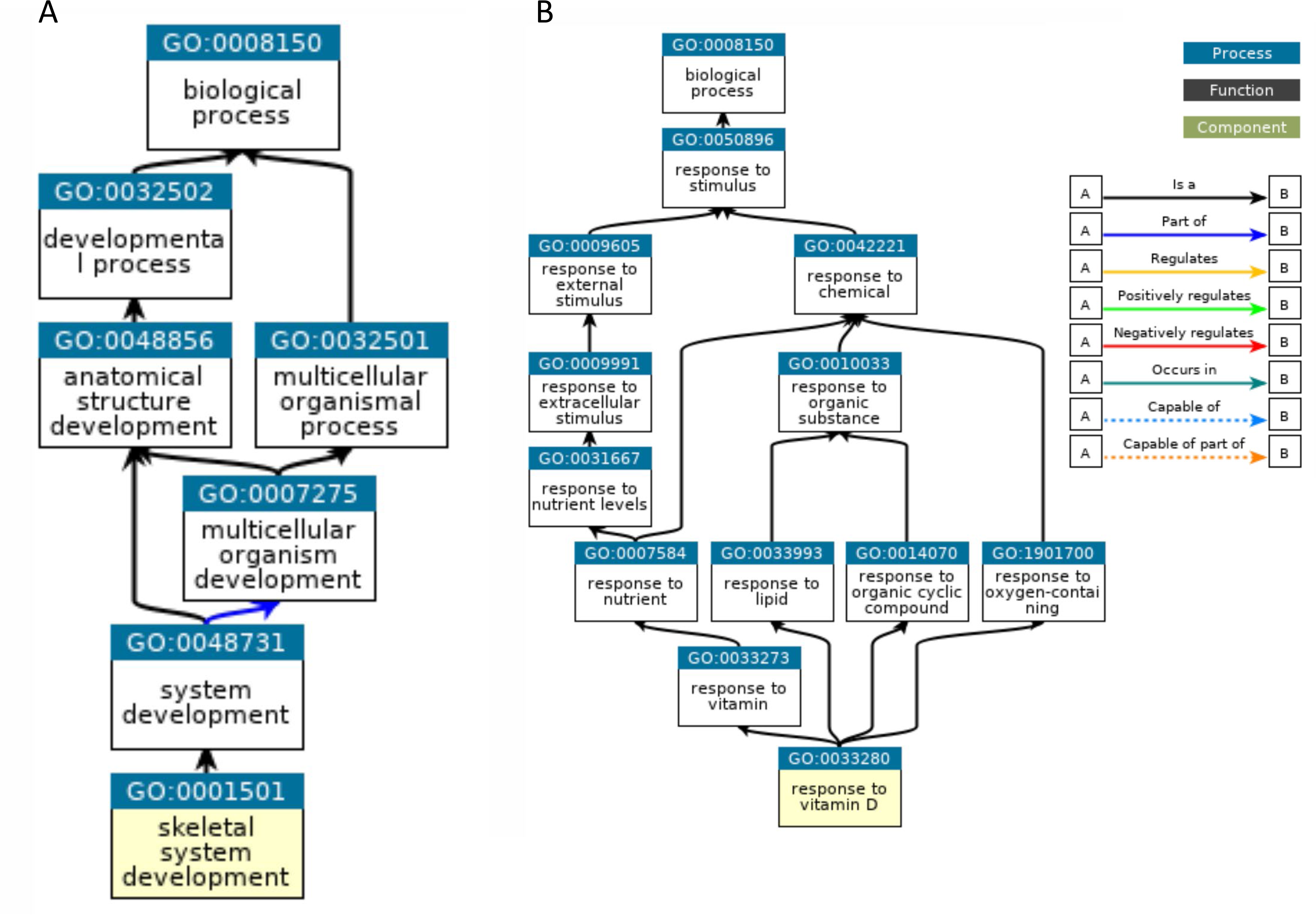
Gene Ontology and Enrichment Analysis. Top GO term identified in GO_Biologic Process for skeletal development and Response to Vitamin D.

**Figure 5.**
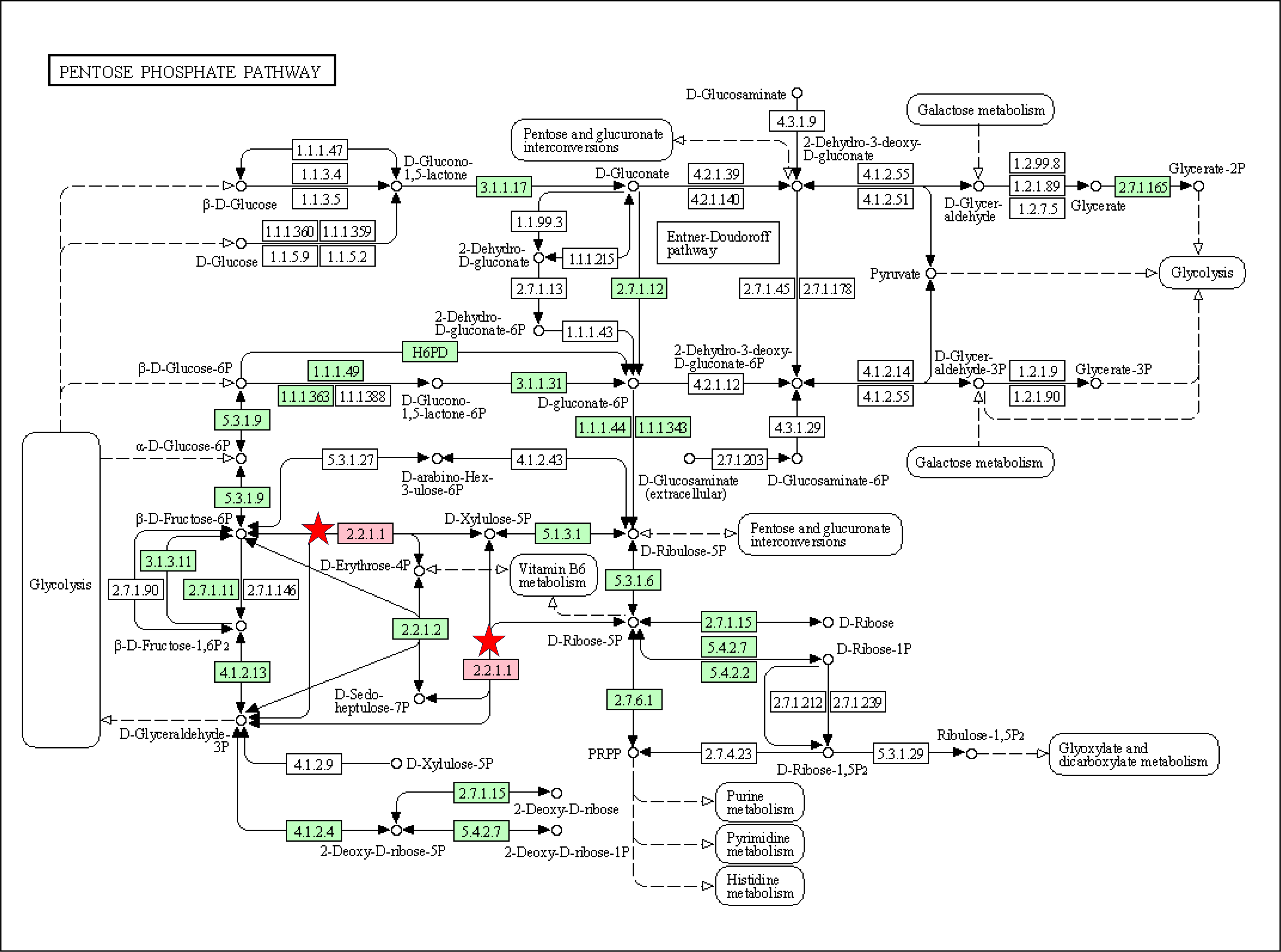
KEGG enrichment analysis flow diagram for pentose phosphate pathway. The red star indicates transketolase enzyme (2.2.2.1) enriched in the MTAG dataset.

**Figure 6.**
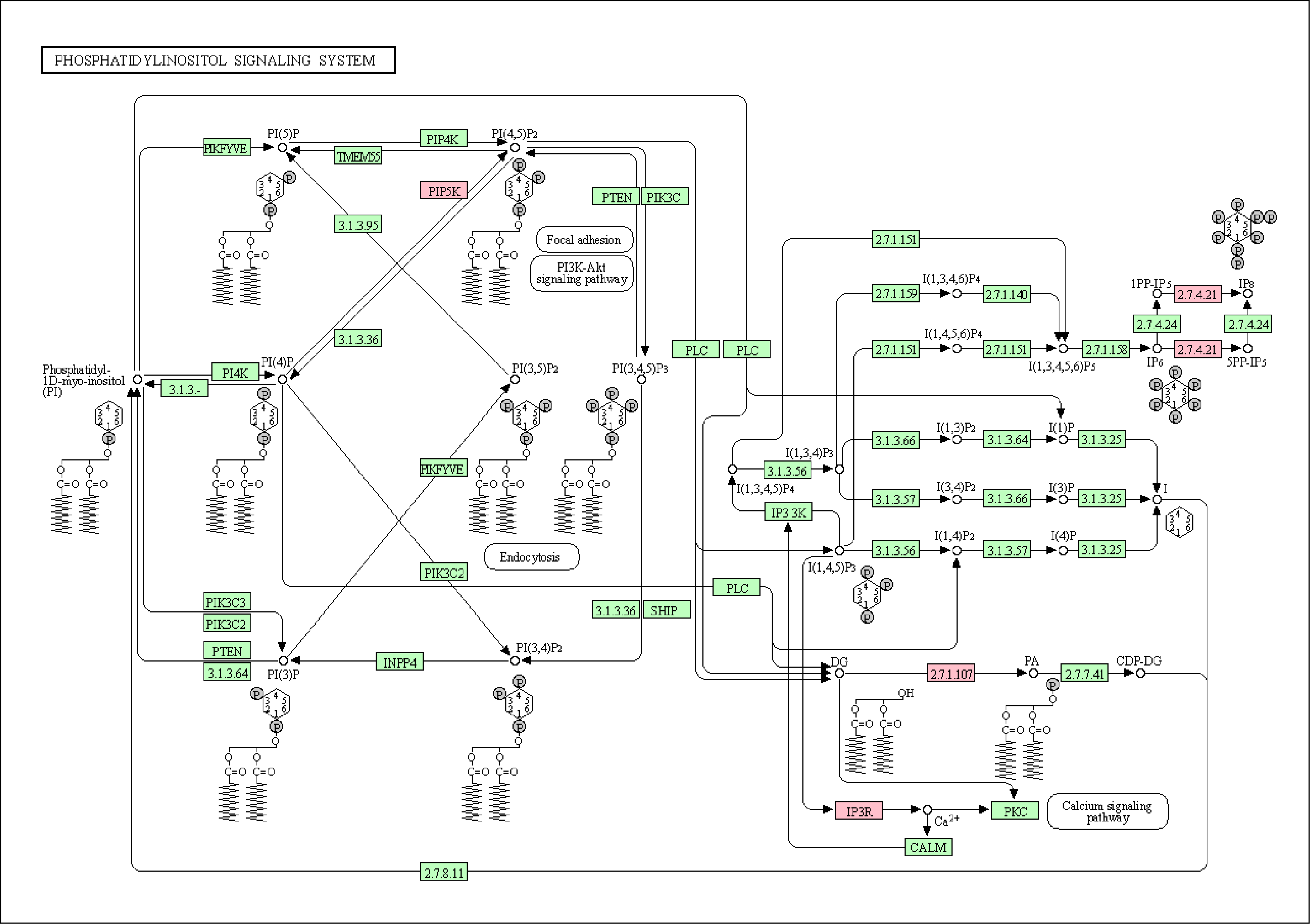
KEGG enrichment analysis flow diagram for phosphatidyl inositol pathway. The pink box indicates genes in the pathway enriched in the MTAG dataset PIP5K, IP3R (ITPR3), 2.7.4.21 (IP6K3), 2.7.1.107 (DGKD)

Secondly, we also found the top SNP, rs73743401, was in proximity to inositol hexakisphosphate kinase 3 *(IP6K3) and* inositol 1,4,5-trisphosphate receptor type 3 *(ITPR3)* genes which are involved in inositol phosphate biosynthesis. *IP6K3* encodes a protein that is involved in AKT signaling. ITPR3 gene encodes a receptor for inositol 1,4,5-trisphosphate, a second messenger that mediates the release of intracellular calcium. Every additional T allele at rs73743401 was associated with ∼ 22% higher FGF23 concentrations in the MTAG analysis (MAF 0.14, Beta coefficient 22.0, P value 5×10^-60^). Thus, functional enrichment analyses identified known links to biological functions of FGF23 and discovered novel pathways linked to pentose phosphate and phosphatidyl inositol signaling pathways.

#### Integrated Network Modeling

Sequence models of disease predicton (eg RNA-seq, GWAS, genome sequencing) can describe the molecular effects of gene mutations and expressions, but the interpretation of how these gene variations leads to disease phenotypes requires understanding of the dysregulated pathways and processes. Using an integrated network modeling approach massive collections of omics data can be summarized to create genome-scale functional maps and find novel gene-interaction networks specific to a biological context. To this end, we utilized a supervised machine learning method that integrates signal from GWAS together with signal in tissue-specific functional networks to reprioritize genes potentially associated with the disease/trait of interest. We found the top canonical pathways were associated with cardiac hypertrophy, multiple sclerosis and hematopoiesis **(Figure 7)**. VDR and TGFβR2 signaling pathways were identified as the top regulator effect networks and MAPK signaling molecules were common to the top 5 causal networks (data not shown). Among the canonical pathways that regulate cell metabolism, pathways for pentose phosphate, phosphatidyl inositol biosynthesis and citrulline-nitric oxide had the highest predicted activation z-scores. Signaling pathway with FGF6, FGF7 and FGF23 had the highest predicted inhibition z-scores ( z-score -2.0, P value 5.4.E-03). We discovered a novel pathway that was previously unreported in the literature involving Coordinated Lysosomal Expression and Regulation (CLEAR) signaling to have the highest predicted activation score ( z-score 2.24, P value 7.7.E-03) **(Figure 7**) and was among the overlapping gene clusters also identified in NetWAS in kidney, bone and heart tissues. CLEAR pathway regulates lysosomal function, and cellular response to nutrient sensing, and 6 genes (*FNIP1, GBA1, GUSB, ITPR3, PPP2R3A, RALB*) in CLEAR pathway were found to have significant z scores **(Tabe 2).** Of note, *ITPR3* in proximity to SNP, rs73743401, was associated with ∼ 22% higher FGF23. Next we queried Ingenuity knowledge database for disease states associated with high activation z-scores for CLEAR pathway and found that 22 genes in this pathway were upregulated in heart tissue obtained from deceased hemodialysis patients with left ventricular hypertrophy (CAIN cohort) and proceeded with an indepth comparison analysis of the two datasets.

**Figure 7.**
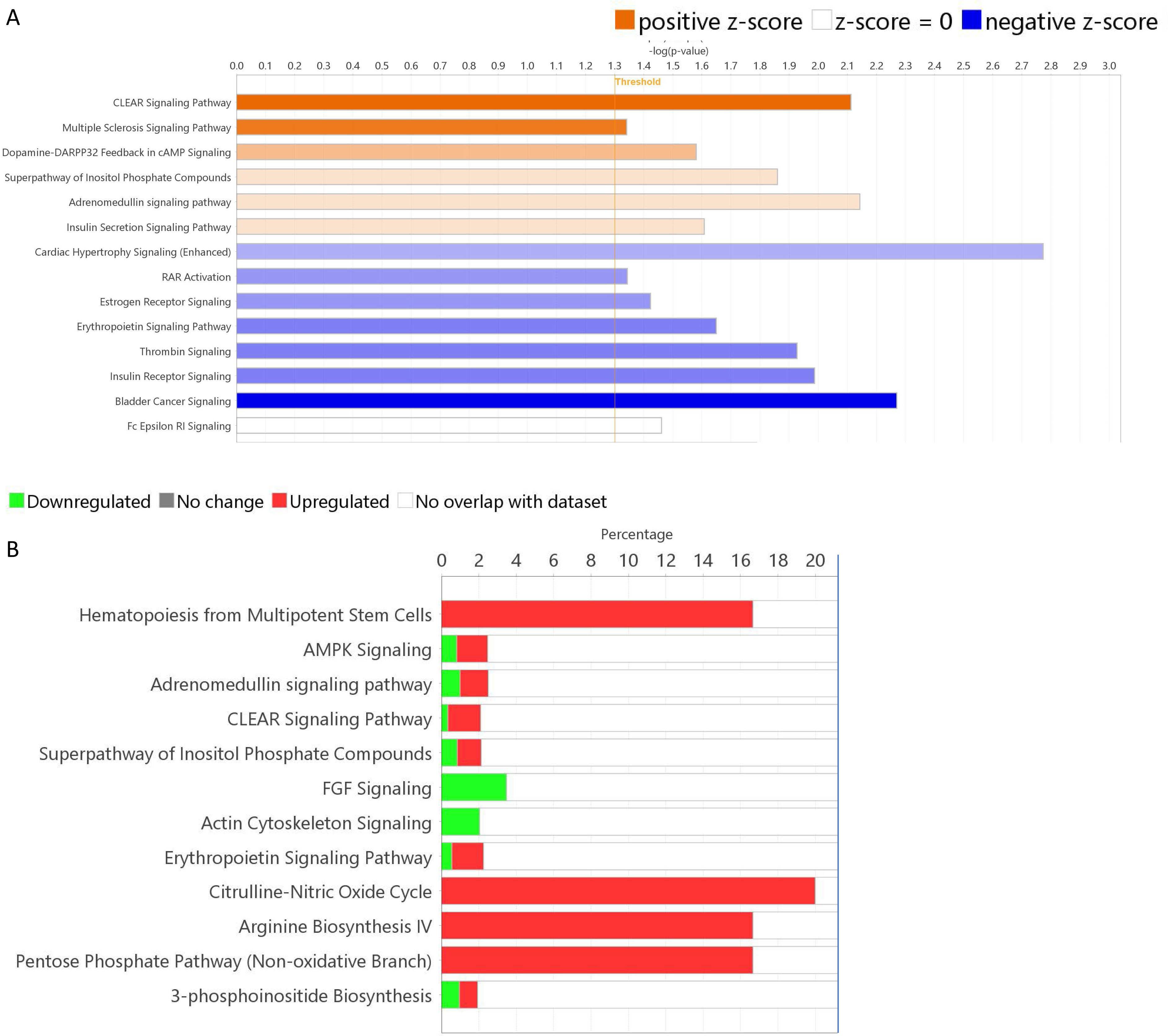
Canonical pathway analysis of MTAG. **A.** Prediction of pathway activation (blue bars) and inhibition (orange bars) represented by z scores. Orange line represents threshold for – log (p-value). **B.** Percent of genes upregulated (red) and downregulated (green) in the enriched canonical pathways shown as percentage.

##### Comparison Analysis of MTAG_FGF23 with cardiac disease in advanced CKD

To further analyze the top signals identified by integrative network modeling, we explored tissue-specific functional networks and patterns in the heart, an organ system that is directly impacted by CKD-MBD and FGF23 excess [23]. We performed comparison analyses of genomic data from MTAG_FGF23 with a publicly available cardiac dataset of CKD. In the CAIN cohort, bulk RNA-seq was performed on left ventricular heart tissue obtained from deceased hemodialysis patients and controls [11]. In this cohort, patients on hemodialysis had significantly larger heart tissue weight and ventricular wall thickness, and greater myocardial fibrosis on histology compared to controls. In the comparison analysis, we found overlapping and distinct pathways; specifically, CLEAR signaling pathway was the top canonical pathway in both datasets with high predicted activation z-scores and multiple sclerosis pathway was predicted to be activated in MTAG_FGF23 but inhibited in left ventricular heart tissue **(Figure 8**). Cardiac hypetrophy and estrogen receptor signaling (aryl hydrocarbon signaling) were common to both datasets. Among the pathways for disease and functions, organismal death had the highest predicted activation score and cell migration had the highest inhibition score in the left ventricle gene expression dataset. We found top genes (KPNA1, PARP9, ITPR3, TKT, Rgs14, FGF23, PTH, DGKD, DNMT3A) in the MTAG dataset from Table 1 overlapped in this network for organismal death from the CAIN cohort **(Figure 9**).

**Figure 8.**
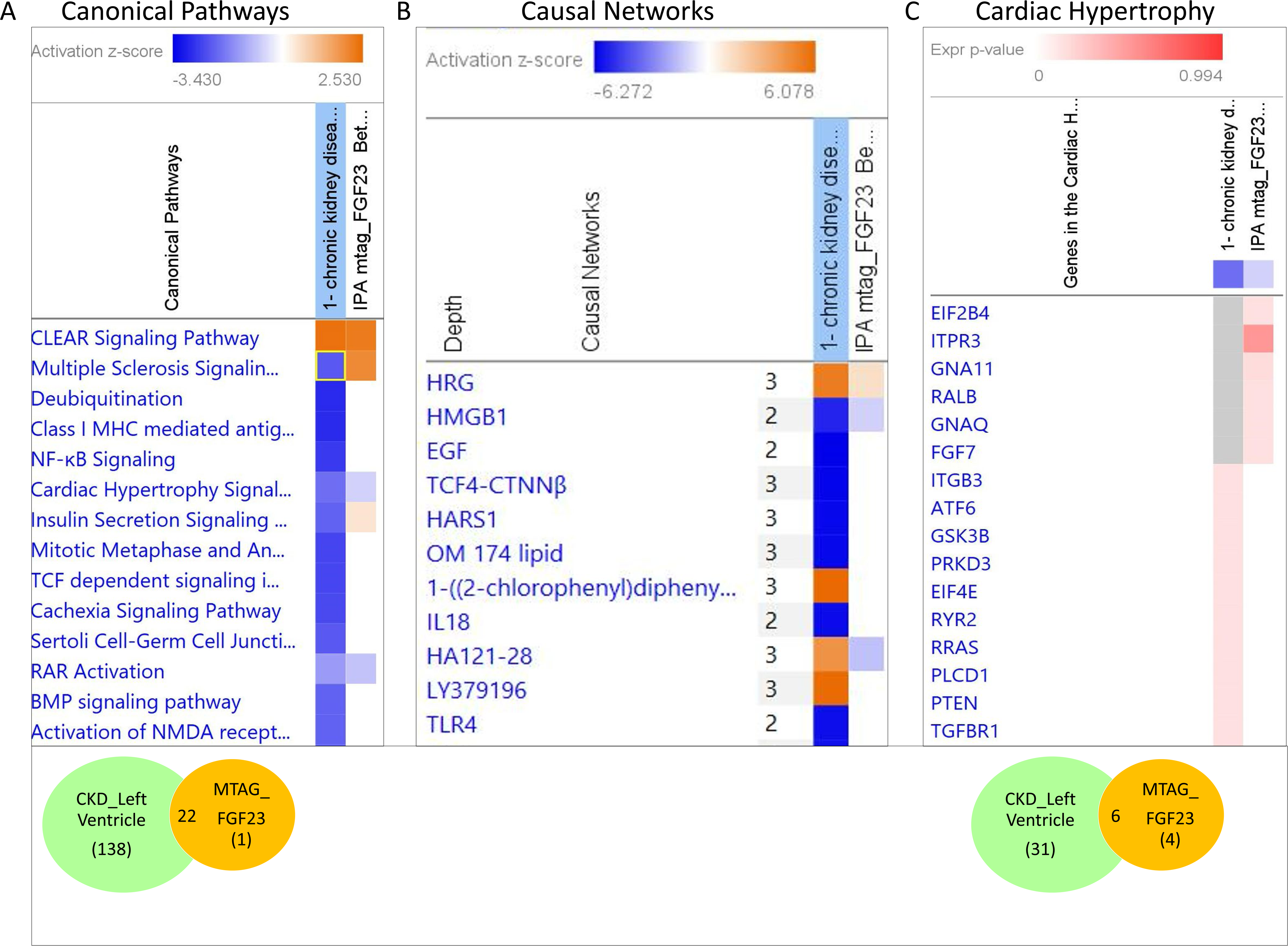
Comparison analysis of lead genetic loci in MTAG with differential gene expression from bulk RNA-seq data of left ventricle of patients with advanced CKD vs controls **A.** Canonical pathways and **B.** Causal networks common and distinct in each dataset. Activation z scores represented as orange boxes for predicted activation and blue boxes for inhibition. **C.** Genes upregulated (orange) and downregulated (blue) or no change (grey) in pathway for cardiac hypertrophy.

**Figure 9.**
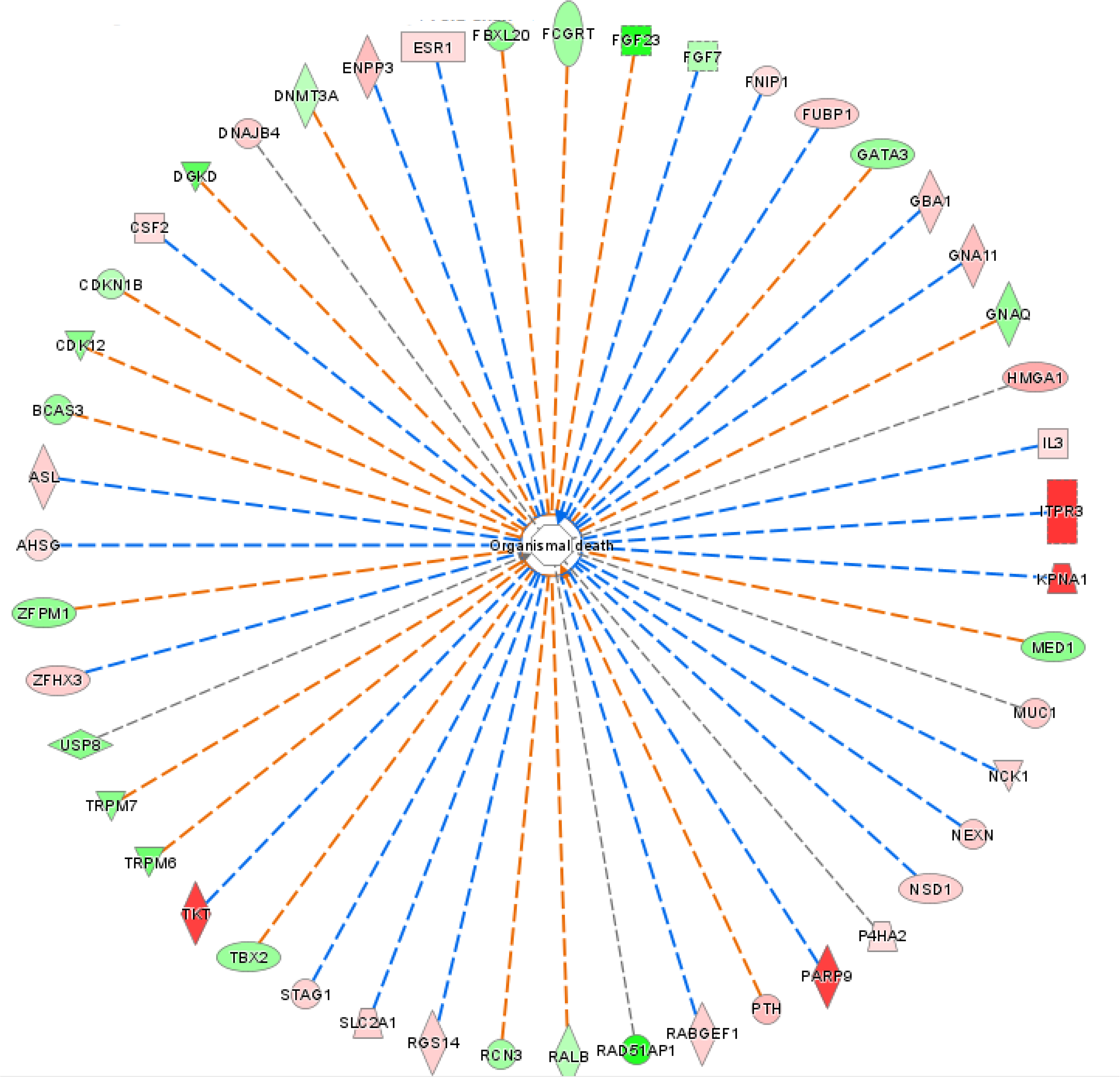
Comparison analysis of lead genetic loci in MTAG with differential gene expression from bulk RNA-seq data of left ventricle of patients with advanced CKD vs controls. Pathway for Organismal death identified in both datasets represented in schematic diagram. Genetic loci are represented in red/pink and in green when associated with higher, or lower, circulating FGF23, respectively in MTAG.

##### HRG-HMGB1 identified as master regulators of downstream canonical pathways linked to both FGF23 and cardiac disease in advanced CKD

We examined the 2 datasets (MTAG_FGF23 and hypertrophied left ventricular heart tissue differential gene expression) for upstream molecules that potentially function as master regulators of disease/trait of interest (ie cardiac diease in CKD). We discovered a novel causal network with histidine-rich glycoprotein and high-mobility group protein box 1 (HRG-HMGB1) as the master regulator of downstream canonical pathways in both datasets. In disease states, HMGB1 is pro-inflammatory and causally associated with renal and cardiovascular injury [24]. Consistent with reported literature, HRG-HMGB1 was upstream of major signaling pathways shown in **Figure 10**. Activation of HRG is predicted to inhibit HMGB1 and downregulate signaling pathways for TGFβ1, TLR, IL-2, IL-6, NFKB, HIFα, and to activate AKT and stat5a/b signaling. In addition, we identified several top genes from Table 1 within this HRG-HMGB1 network as shown in **Figure 10**. Specifically, FAM162a, TKT, PARP9 and DTX3L genes were activated and FGF23, ST3GAP, USP8, BCAS3, GNAQ, PCGRT and PPP1R1B were inhibited by HRG-HMGB1 signaling networks.

**Figure 10.**
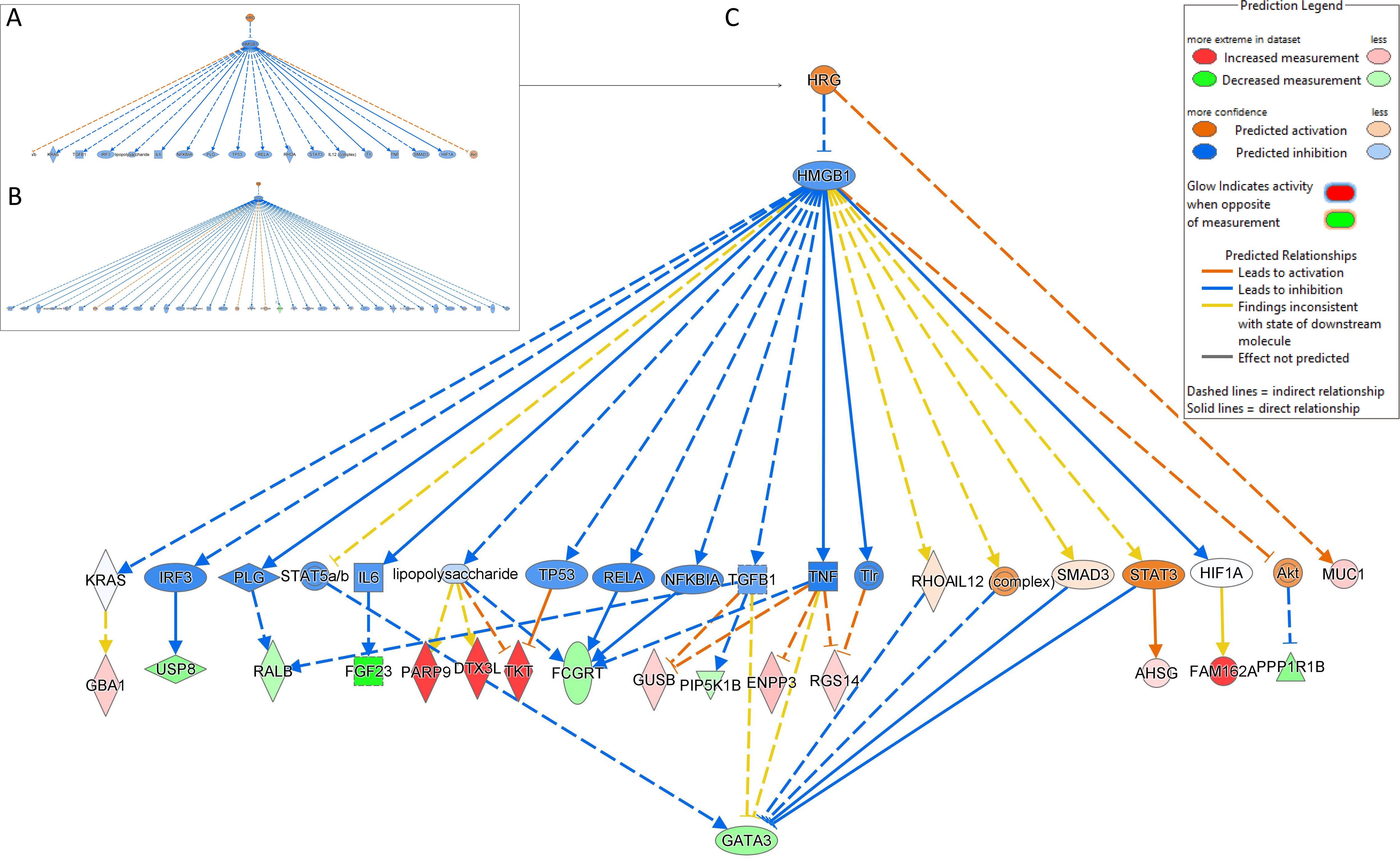
Comparison analysis of lead genetic loci in MTAG with differential gene expression from bulk RNA-seq data of left ventricle of patients with advanced CKD vs controls. Causal network analysis revealed HRG-HMGB1 as a master regulator for downstream canonical pathways in **A.** and **C.** MTAG FGF23 dataset, and **B.** CKD Left ventricle dataset.

## DISCUSSION

Computational biology has revolutionized our capabilities to understand complex pathophysiology such as CKD [25]. We leveraged a comprehensive list of computational and deep machine learning tools to expand networks and pathways linked to genetic traits associated with CKD-MBD. In this study we used a novel methodology, MTAG, to analyze GWAS from multiple large consortiums, to boost the number of genome-wide significant loci for FGF23. We then used a multi-pronged functional genomics approach to model interactive and dynamic networks to discover novel associations between these genetic traits and FGF23 signaling. Using NetWAS, we identified tissue-specific gene clusters in kidney, bone and heart including modules that regulate mineral ion homeostasis. Using integrative network modeling, we identified known (VDR,TGFβR2 and MAPK) and novel (CLEAR, pentose phosphate, phosphatidyl inositol biosynthesis) pathways linked to FGF23. Lastly, we explored tissue-specific functional networks and patterns to discover novel genetic traits (CLEAR, HRG-HMGB1, cardiac hypetrophy and estrogen receptor signaling) that overlapped between MTAG_FGF23 and in cardiac tissue of deceased patients with advanced CKD. Thus, combining clinical and experimental data with insilico analysis we have advanced our understanding of genetic drivers of CKD-MBD and shed light on potential novel mechanisms that mediate cardiovascular disease in CKD.

MTAG was developed to boost the statistical power of GWAS by incorporating information from effect estimates across traits. Advantages of MTAG over other analyses include unique capability to combine analysis of multiple, genetically correlated traits (e.g. Ca, P, vit D, PTH and FGF23) using summary statistics of GWAS rather than requiring individual genotypes. MTAG mitigates bias from sample overlap which is often present when using discovery samples for different traits to calculate linkage disequilibrium (LD) score regression [10]. With this approach we identified novel genetic traits previously unreported to be associated with circulating FGF23 (Table 1).

Given that CKD-MBD is a complex disease process. we utilized multiple analyticial tools to understand how genetic traits identified with MTAG align with biological networks in a tissue- and cell-specific manner in the context of FGF23. First, we used NetWAS analyses to minimize bias from “missing heritability” to identify the cumulative weak effects of variants and to minimize literature bias where candidate genes not found in the literature but exhibit strong support for pathogenesis can be discovered agnostically [16, 18]. Throught this approach. we built gene clusters and modules for 3 organ systems; kidney, bone and heart, that are intricately involved in CKD-MBD. Some of the gene modules were distinct (i.e tissue-specific) whereas others overlapped across organ systems suggesting that similar pathways are involved in pathogenesis of CKD-MBD in the kidney, bone and heart. Notably, we found aryl hydrocarbon signaling pathway common to all 3 organ systems and transcription factor, HNF4α, was identified as a top upstream regulator. HNF4α2 isoform was recently implicated in the pathogenesis of renal osteosystrophy and plays a critical role in cell death and osteogenesis [26]. HIF1α, TGFβ, mitochondrial dysfunction, Nrf2-mediated oxidative stress response and PPAR α/RXR α were also among the top signaling pathways common to all 3 tissues of interest.

Using a series of enrichment analysis and network modeling we discovered known (skeletal development and vitamin D) pathways associated with FGF23 confirming the SNPs in the MTAG dataset are biologically relevant. We also found novel pathways including pentose phosphate pathway and phosphatidyl inositol signaling were highly enriched in both KEGG and IPA analyses. The top SNP, rs73188394, was in proximity to *TKT* gene that belongs to pentose phosphate pathway, and, rs73743401, was in proximity to *IP6K3 and ITPR3* genes that belongs to the phosphatidyl inositol signaling and CLEAR pathway. Both SNPS were associated with 21-22% higher FGF23 concentrations in the GWAS consortium. TKT encodes for transketolase enzyme that plays a critical role in energy metabolism and oxidative stress response. Little is known about role of TKT in CKD but pentose phosphate pathway is high conserved in evolutionary biology [22]. Iron, calcium and phosphate are required to support non enzymatic reactions in the pentose phosphate pathway which also provides substrates for glycolysis, fatty acid, steroid, DNA and RNA synthesis. Recently, it was reported that kidney-specific glycolysis pathway serves as a phosphate sensor and its by product, glycerol-3-phosphate (G-3-P) released into circulation upregulates FGF23 production in bone [27]. Given that pentose phosphate pathway is tightly linked to glycolysis by providing substrates and energy via NADPH, TKT may play a central role connecting FGF23 and MM to vital cellular functions including glucose, lipid, energy metabolism, renal tubular phosphate transport and response to hypoxia.

Our analyses identified CLEAR signaling pathway as a key causal network with the highest predicted activation z-scores and 6 genes (*FNIP1, GBA1, GUSB, ITPR3, PPP2R3A, RALB*) within this pathway belonged to our dataset. In addition, we found 22 genes in CLEAR pathway were upregulated in cardiac tissue obtained from deceased hemodialysis patients (CAIN cohort) and thus proceeded with an in depth comparison analysis of the 2 genomic datasets. In the CAIN cohort, cause of death was not reported but histology confirmed the diagnosis of left ventricular hypertrophy and fibrosis in hemodialysis patients compared to controls [11]. Gene expression data revealed that CLEAR signaling pathway was the top canonical pathway with high predicted activation z-scores in the CAIN dataset. Cardiac hypetrophy and estrogen receptor signaling pathway activation were also common to both datasets. Among the pathways for disease and functions, organismal death had the highest predicted activation score in the left ventricle gene expression dataset which is expected since the patients died on dialysis. Interestingly, we found several genes (KPNA1, PARP9, ITPR3, TKT, Rgs14, FGF23, PTH, DGKD, DNMT3A,) in the MTAG dataset were present in this network for organismal death from the CAIN cohort. Notably, FGF23 and PTH are associated with cardiovascular disease and mortality in patients with CKD [28–30]. FGF23 directly induces left ventricular hypertrophy via FGFR activation [23, 31] and induces cardiac fibrosis via TGFβ signaling pathway [32]. Recently, DNMT3a was shown to play an important role in kidney development and DNMT3a-induced methylation of specific DNA regions was found to be enriched in kidney disease risk loci in GWAS studies of humans with CKD. This suggests that DNMT3a plays a key role in epigenitic modification that contributes to CKD progression [33]. Through GWAS studies of circulating mineral markers in the general population, we and others have shown that rs4074995 (RGS14) SNP was the top SNP associated with circulating PTH, calcium, phosphate, and FGF23 [2–5, 34]. Moreover, in adults with CKD, the minor allele of rs4074995 (RGS14) was associated with lower phosphorus and FGF23 levels and lower prevalence of hyperparathyroidism [3]. We performed expression quantitative trait loci analysis (eQTL) and showed that rs4074995 was associated with lower RGS14 gene expression in the kidneys and colocalization studies demonstrated strong correlations between RGS14 gene expression and circulating FGF23 and PTH [3]. These findings align with recent preclinical studies that provide evidence for RGS14 to play a key role in convergence of FGF23- and PTH-dependent signaling in the renal proximal tubule to inhibit phosphate reabsorption via sodium phosphate co transporter, Npt2a [35, 36]. Further studies are needed to understand the role of Rgs14 in cardiovascular disease in CKD.

Another key finding from our analysis was the identification of HRG-HMGB1 as the master regulator of downstream canonical pathways in both datasets. Activation of HRG is predicted to inhibit HMGB1 and regulates several downstream signaling pathways including TGFβ1, TLR, IL-2, IL-6, NFKB, HIF1α and to activate AKT and stat5a/b signaling. We found FAM162a, TKT, PARP9 and DTX3L genes are activated and FGF23, ST3GAP, USP8, BCAS3, GNAQ, PCGRT and PPP1R1B are inhibited by HRG-HMG signaling networks. HMGB1 is a member of the high-mobility group proteins that are secreted and can be detected in blood and urine [24]. HMGB1 is ubiquitously expressed and is involved in cellular damage and repair. The biological activity of HMGB1 depends on its subcellular localization. In the nucleus, HMGB1 regulates DNA repair chromatin remodeling and maintains nucleosome and telomere homeostasis. In the cytoplasm, HMGB1 regulates autophagy and mitchondrial function to regulate cell death. Extracellular HMGB1 acts as a damage-associated molecular pattern (DAMPs)/alarmin to promote cell migration and proliferation but one of its key functions is to accelerate cell death by inducing lysosomal membrane permeabilization. In a cohort of non diabetic CKD, serum HMGB1 was significantly elevated and independently correlated with asymmetric dimethylarginine (ADMA), a marker of endothelial dysfunction and cardiovascular diseases, indicating that HMGB1 is actively involved in CKD progression and might lead to the development and progression of cardiovascular disease [37]. In our study, we show that HRG-HMGB1 complex is a master regulator of canonical pathways in patients with advanced CKD and LVH and the same pathways are associated with FGF23 signaling networks. Future studies will investigate the precise role for these novel genetic traits in the pathophysiology of cardiovascular disease in CKD and FGF23 excess.

In conclusion, our study has shed light on key genetic drivers of CKD-MBD, identifiying novel SNPs associated with circulating FGF23 and with the aid of computational tools and machine learning we have generated a catalog of established and novel networks and pathways that link FGF23 to complications of CKD.

## Data Availability

All data produced in the present study are available upon reasonable request to the authors

https://www.ukbiobank.ac.uk/

https://web.chargeconsortium.com/

https://www.ncbi.nlm.nih.gov/geo/

## AUTHOR CONTRIBUTIONS

FP and CRC conceived the study and designed the experiments; EA and NV performed experiments; FP, EA, CRC, PAF, LJS analyzed and interpreted data; FP EA and CRC wrote the manuscript and all authors approved and commented on the manuscript.

## ACKNOWLEDGMENTS

This work was supported by National Institutes of Health (NIH) R01DK122075 (CRC). The funders had no role in study design, data collection and interpretation, or the decision to submit the work for publication.

